# Assessing The Feasibility of AI-Driven Systems for Early Detection of Infectious Diseases at Julius Nyerere International Airport, Tanzania: Policy, Infrastructure, and Ethical Considerations

**DOI:** 10.64898/2026.04.08.26350459

**Authors:** Elizabeth Ezekiel Malingumu, Isimbi Badaga, Dotto Daniel Kisendi, Rasabs Wende Pierre Kabore, Onacis Guerde Yeremon, Mohamed Ally Mohamed, Qun He

**Affiliations:** Department of Preventive Services, Section of Environmental Health, Hygiene and Sanitation, Ministry of Health, Dodoma, Tanzania; Rwanda Biomedical Centre, Rwanda; National Health Insurance Fund, Directorate of Membership Services, Morogoro, Ministry of Health, Tanzania; Dori Health District, Ministry of Health, Sahel region, Bukina Faso; Outpatient Care Center of the Bangui Community Hospital, Ministry of Health, Central African Republic; Health System Strengthening, East, Central, and Southern Africa Health Community (ECSA HC), Arusha, Tanzania; Dermatology Hospital of Southern Medical University, China

## Abstract

This study evaluates the feasibility of implementing artificial intelligence (AI)-driven disease surveillance systems at Julius Nyerere International Airport (JNIA) in Tanzania, a key hub for regional and international travel. Through a mixed-methods approach combining qualitative interviews and quantitative surveys, the research assesses the infrastructure, human resource capacity, and regulatory frameworks necessary for AI integration. Findings indicate that while Port Health Officers are strongly optimistic about AI’s potential to enhance disease detection, the airport faces significant barriers, including outdated infrastructure, insufficient technical resources, and a lack of trained personnel. Ethical and privacy concerns, particularly surrounding data security, also emerged as key challenges, compounded by limited public awareness and the socio-cultural acceptability of AI systems. Furthermore, the study identifies gaps in national policies and inter-agency coordination that hinder the effective implementation of AI technologies. The research concludes that while current conditions render AI adoption infeasible, strategic investments in infrastructure, workforce training, and policy development could pave the way for future integration, enhancing public health surveillance at JNIA and potentially other airports in low- and middle-income countries. This study contributes critical insights into the barriers and opportunities for AI-driven disease surveillance in low-resource settings, specifically focusing on a high-priority transit point, international airports. It emphasizes the importance of region-specific solutions to enhance health security in East Africa and supports the broader global health agenda by advocating for international collaboration and the development of scalable disease surveillance systems. Future research should explore pilot AI implementations at other airports to evaluate real-world challenges and refine AI systems for broader applicability, including cost-effectiveness analyses and integration of public perspectives on AI.

## 1. Introduction

The rapid advancement of artificial intelligence (AI) has opened new avenues in healthcare, offering innovative solutions to long-standing challenges in disease detection, monitoring, and prevention. AI empowers machines to simulate human cognitive functions, such as learning and problem-solving, to enhance medical practice and healthcare delivery(1, 2). In the realm of infectious disease surveillance, AI has shown significant promise in improving early detection, response times, and predictive capabilities, critical factors in managing the spread of infectious diseases(3).However, the application of AI-driven systems in disease surveillance at international airports, particularly in Tanzania, remains underexplored.

International airports are vital nodes in the global network of travel, serving as major points of entry for passengers and, consequently, for the spread of infectious diseases(4). While airports like Julius Nyerere International Airport (JNIA) are essential to regional and international connectivity, they also face unique challenges in managing health risks, given the high volume of travelers and the often-limited capacity of disease surveillance systems. Traditional disease monitoring techniques, which rely primarily on manual screenings and reactive measures, are insufficient for handling the rapid transmission of infectious diseases, particularly in high-traffic environments(5). The integration of AI-driven systems could revolutionize disease detection at airports, offering real-time, automated surveillance that significantly enhances early identification and response to emerging health threats(6). However, the feasibility of implementing such systems at JNIA has not been comprehensively investigated.

Tanzania, like many countries in East Africa, faces substantial public health challenges, including high rates of malaria, tuberculosis, and cholera. The country’s healthcare infrastructure is often under-resourced, which compounds the difficulty of implementing efficient disease surveillance systems(7). The growing volume of air traffic at JNIA underscores the need for advanced disease-detection and monitoring technologies to prevent the spread of infectious diseases. AI presents an opportunity to address the gaps in the current surveillance system by providing more accurate, integrated, and proactive solutions(8).

This study aims to assess the feasibility of implementing AI-driven systems for early disease detection at JNIA. Specifically, it will examine the airport’s infrastructure, identify capacity gaps, explore ethical implications, and evaluate the regulatory frameworks necessary to support the integration of AI technologies. By focusing on JNIA, this research will provide valuable insights into the challenges and opportunities of adopting AI solutions in disease surveillance, offering a model for other airports in Tanzania and potentially other low- and middle-income countries. Ultimately, this study seeks to enhance public health preparedness at one of Tanzania’s most critical international transit points, thereby improving disease surveillance and response capabilities in the region.

## 2. Materials and Methods

### 2.1. Research Design

This study employed an exploratory sequential mixed-methods design, integrating qualitative and quantitative components. The qualitative phase used semi-structured interviews guided by the Consolidated Framework for Implementation Research (CFIR) to explore barriers, facilitators, governance structures, and ethical considerations for AI integration. The quantitative phase utilized structured surveys and a standardized infrastructure checklist to assess current surveillance operations, as well as to evaluate AI-compatible infrastructure such as internet connectivity, data storage, power reliability, and dedicated technical facilities at JNIA.

### 2.2. Study area

The study was conducted at Julius Nyerere International Airport (JNIA), Dar es Salaam, Tanzania. The country’s primary international aviation hub. JNIA handles approximately 2.8 million passengers annually and serves as a critical port of entry for regional and international travel, making it a high-priority site for infectious disease surveillance and cross-border health security. The airport maintains formal port health services aligned with the International Health Regulations (IHR 2005), with a clearly defined organizational structure and designated public health personnel responsible for disease screening, monitoring, and response.

**Figure 1:**
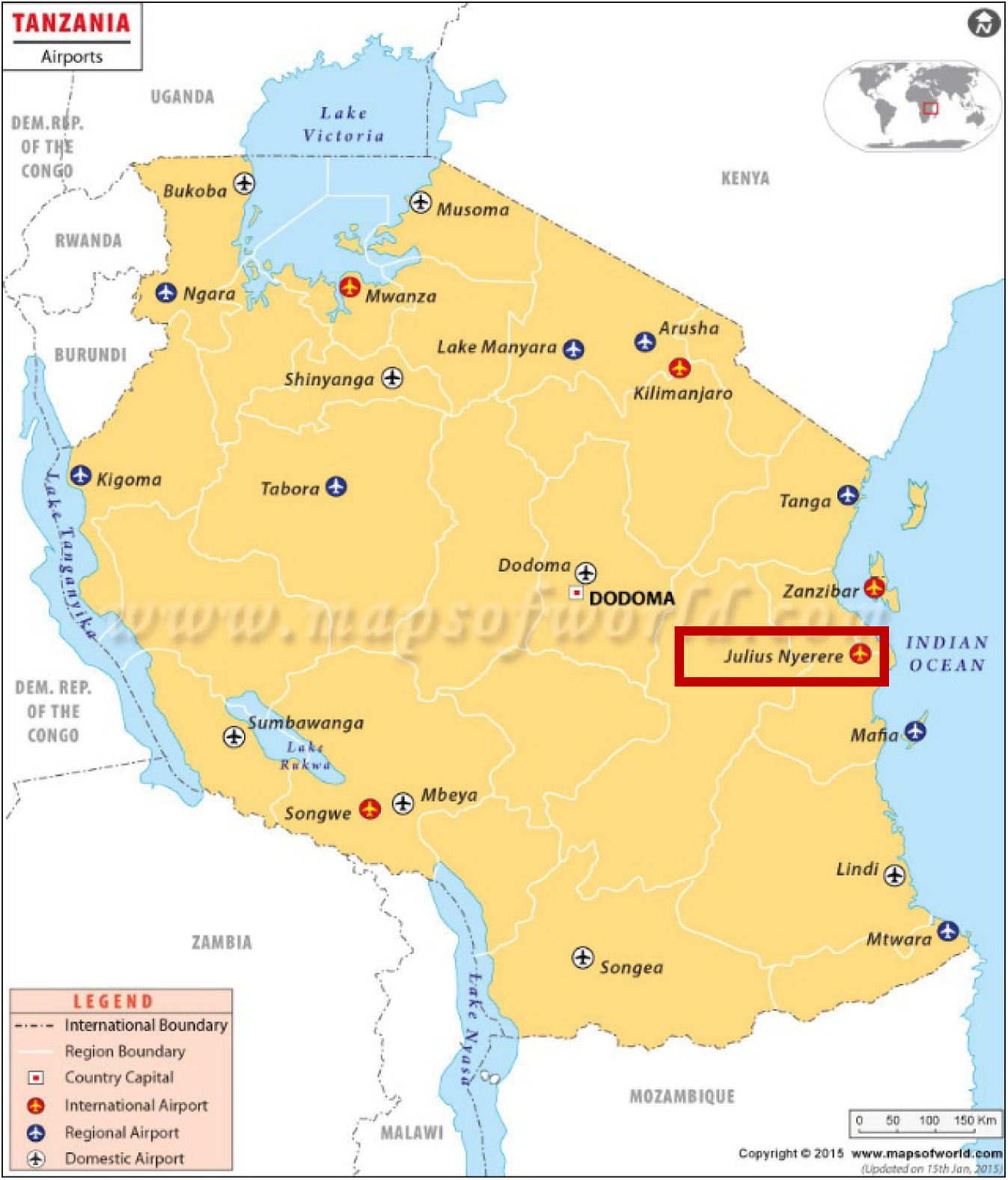
Map showing Airports in Tanzania

### 2.3. Study Population and Sampling

#### 2.3.1. Qualitative Component

The qualitative sample comprised key stakeholders directly involved in port health, airport management, and ICT governance at JNIA. A purposive sampling strategy was used to select participants with substantial professional experience and direct involvement in disease surveillance operations. Inclusion criteria were: direct involvement in port health surveillance, airport operations, or health-related ICT management at JNIA, a minimum of 3 years of relevant professional experience, practical experience implementing public health screening or surveillance protocols at the airport, and relevant roles in policy, governance, or technical oversight of airport health systems. A total of 8 participants were interviewed, including 2 officials from the Tanzanian Ministry of Health, 2 port health officers stationed at JNIA, 2 representatives from the Tanzania Airports Authority, 1 airport management officer, and 1 airport-based IT specialist Sample size was determined by information saturation and the principle of information power. No new themes, codes, or substantive insights emerged after the eighth interview, confirming that the data were sufficiently rich and comprehensive to address the study objectives.

#### 2.3.2. Quantitative Component

The quantitative component targeted all Port Health Officers (PHOs) involved in implementing disease surveillance and screening measures at JNIA. Participant recruitment took place from 6^th^ to 30^th^ August 2025. A census sampling method was employed, inviting participation from all 43 active PHOs at the airport. However, only 26 PHOs participated in the survey. Despite efforts to engage all PHOs, some officers were unable or unwilling to participate due to the demands of their daily responsibilities. Additionally, some officers were reluctant to participate due to privacy concerns. To mitigate these challenges, follow-up reminders were sent to non-respondents, and officers on leave were given the opportunity to participate once they returned to duty. Despite these efforts, not all officers could be reached or persuaded to participate in the study. Nevertheless, the 26 PHOs who participated were considered to represent a comprehensive range of perspectives, providing valuable insights into the operational and infrastructural challenges related to disease surveillance at JNIA. The research team, in collaboration with JNIA’s IT and port health departments, completed an infrastructure checklist to ensure objective, standardized data collection.

### 2.4. Data Collection Methods

#### 2.4.1. Qualitative Data Collection

Qualitative data were collected via semi-structured interviews, conducted either face-to-face or by telephone, based on participant availability. All interviews were facilitated by a single researcher to ensure consistency in questioning and data capture. Each interview lasted 30–40 minutes, with detailed notes taken during each session, followed by an immediate post-interview review to verify accuracy, clarify ambiguities, and ensure completeness of data capture. The interview guide was structured around the five core domains of the CFIR framework to systematically explore barriers and enablers of AI implementation. The guide was pilot-tested with two PHOs not included in the final sample to refine question clarity, cultural appropriateness, and contextual relevance.

#### 2.4.2. Quantitative Data Collection

##### Structured survey

A structured online survey was created using Google Forms and shared through the official JNIA PHO WhatsApp group. The survey evaluated current surveillance methods, screening processes, response times, equipment availability, and perceived challenges to automation and digitalization. The survey tool was pilot-tested with 5 PHOs to improve question logic, reduce ambiguity, and ensure alignment with JNIA’s operational context. Quality control steps included pre-survey testing with port health staff to enhance question clarity and logic, real-time survey monitoring and follow-up reminders, built-in data validation within the online form (such as mandatory responses for key questions and range checks for numerical data) to decrease response errors, and secure, anonymized storage of responses with access limited to the research team to ensure data confidentiality and ethical compliance.

##### Infrastructure checklist

A standardized infrastructure checklist was used to assess the availability, functionality, and adequacy of physical and digital resources required for AI system deployment at JNIA. Domains assessed included network connectivity, computing hardware, data storage, power supply and backup, dedicated workspace, and system interoperability. The checklist was developed based on published guidelines for AI implementation in public health settings and validated by an external expert in digital health infrastructure, ensuring its scientific rigor and relevance to the study’s objectives.

### 2.5. Data Analysis

#### 2.5.1. Qualitative Analysis

Qualitative data were transcribed verbatim, coded, and thematically analyzed using NVivo 14 Pro. A hybrid deductive–inductive thematic approach was applied, using a pre-defined codebook aligned with the study’s three core research themes: Infrastructure and human resource capacity, Ethical considerations and contextual adaptability, and Policy, regulatory frameworks, and governance. The inductive component allowed for the identification of emergent themes, a methodologically sound practice for exploratory research. A thematic saturation tracking matrix was used to monitor data completeness; saturation was achieved at the eighth interview, confirming that the sample size was sufficient to address the study’s objectives. Methodological rigor was ensured through adherence to the COREQ (Consolidated Criteria for Reporting Qualitative Studies) checklist, a gold standard for reporting qualitative research in high-impact journals.

#### 2.5.2. Quantitative Analysis

Quantitative data from surveys and the infrastructure checklist were analyzed using descriptive statistics (frequencies, percentages, means) to summarize key findings.

The infrastructure readiness for AI-driven disease surveillance at JNIA was assessed using a standardized checklist of 17 core components, comparing available quantities against required quantities for optimal AI operation. Readiness was calculated using the following formula:

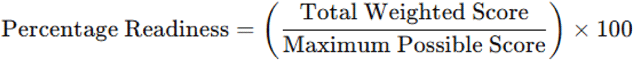

### 2.6. Validity and Reliability

The study’s validity was enhanced through data triangulation, combining qualitative interviews, structured surveys, and infrastructure assessments. Peer debriefing, member checking, and pilot testing of instruments further strengthened the reliability of the findings. External audits of the research process were conducted to ensure rigor and transparency.

Reliability was ensured through clear documentation of all research procedures and the use of external audits. The test-retest reliability of the survey instrument was assessed by having a subset of participants complete the survey again after a specified period. The consistency of data collection was further validated through pilot testing and careful monitoring during the survey administration.

### 2.7. Ethical Considerations

Ethical approval was obtained from Southern Medical University and the Airport Authority in Tanzania (Approval Number: GA/32/469/01/). All participants provided written or electronic informed consent after receiving clear information regarding the study’s purpose, procedures, confidentiality, and the right to withdraw without penalty or negative consequences.

All data were anonymized, with no personal identifiers (e.g., names, job titles) used in analyses. Data were stored securely on password-protected servers, with access restricted to the research team. To address concerns about sensitivity raised by some PHOs, all survey responses were aggregated in analyses to prevent the identification of individual participants or their perspectives.

## 3. Results

### 3.1. Sociodemographic Characteristics of Quantitative Respondents

The quantitative survey was conducted with 26 out of 43 active Port Health Officers (PHOs) at Julius Nyerere International Airport (JNIA), resulting in a response rate of 60%. The sociodemographic characteristics of the respondents are summarized in Table 1. The JNIA PHO workforce is predominantly male (68%) and has a relatively young composition: 67% of respondents are aged 25–45 years, with the average age being 40 years. The workforce is evenly divided between junior (53.85%) and senior (46.15%) PHOs, indicating a balanced mix of experience levels. Regarding tenure, the average duration of employment at JNIA is 5 years, with 34.62% having 5–10 years of experience and 11.54% with over 10 years, reflecting a relatively experienced team. Only 15% of PHOs have less than one year of experience, suggesting stability within the port health team.

**Table 1:** Sociodemographic characteristics of quantitative respondents (n= 26)

| Characteristics | Frequency (N) | Percentage (%) | Mean |
| --- | --- | --- | --- |
| Gender Distribution |  |  |  |
| Male | 18 | 68 |  |
| Female | 8 | 32 |  |
| Age Distribution |  |  |  |
| 25–45 | 17 | 6 |  |
| 45 and above | 9 | 33 |  |
|  |  |  | 40 |
| Current Role (Port Health Officers) |  |  |  |
| Junior Port Health Officer | 14 | 53.85 |  |
| Senior Port Health Officer | 12 | 46.15 |  |
| Duration of Work at Airport (Years) |  |  |  |
| Less than 1 Year | 4 | 15 |  |
| 1–4 Years | 10 | 38.46 |  |
| 5–10 Years | 9 | 34.62 |  |
| 10+ Years | 3 | 11.54 |  |
|  |  |  | 5 |

### 3.2 Current Status of Disease Surveillance and AI Integration at JNIA

Quantitative analysis of survey responses from 26 JNIA PHOs revealed key insights into current disease surveillance practices, perceptions of AI integration, and operational challenges. Table 2 summarizes the findings related to staffing, AI perceptions, screening practices, and recommended improvements. Key findings from the quantitative survey include; 43% of PHOs reported sufficient staffing levels, indicating that nearly 60% perceive a potential staffing gap in port health operations at JNIA. The majority of Port Health Officers (73%) believe that AI could enhance surveillance capabilities, and 73% also felt that AI could improve detection accuracy. Regarding the average number of passengers screened daily, 77% of JNIA officers reported screening between 1000-5000 passengers, with an average of 3000 passengers screened per day. The average screening-to-management time was reported to be 1.5 hours, indicating a slightly more time-intensive process. All officers at JNIA reported that response times to disease detection and action were immediate, with actions taken within 10 minutes of detection. 80% of PHOs rated current screening technology as effective or very effective, with thermoscanners identified as the most commonly used screening tool. However, challenges to AI integration were identified, including a lack of technological infrastructure (65%), high costs (55%), and a lack of skilled workforce (50%). The majority of respondents recommended several improvements for AI integration: 90% suggested enhanced staff training, 85% recommended investments in technology, and 80% called for increased funding.

**Table 2:**
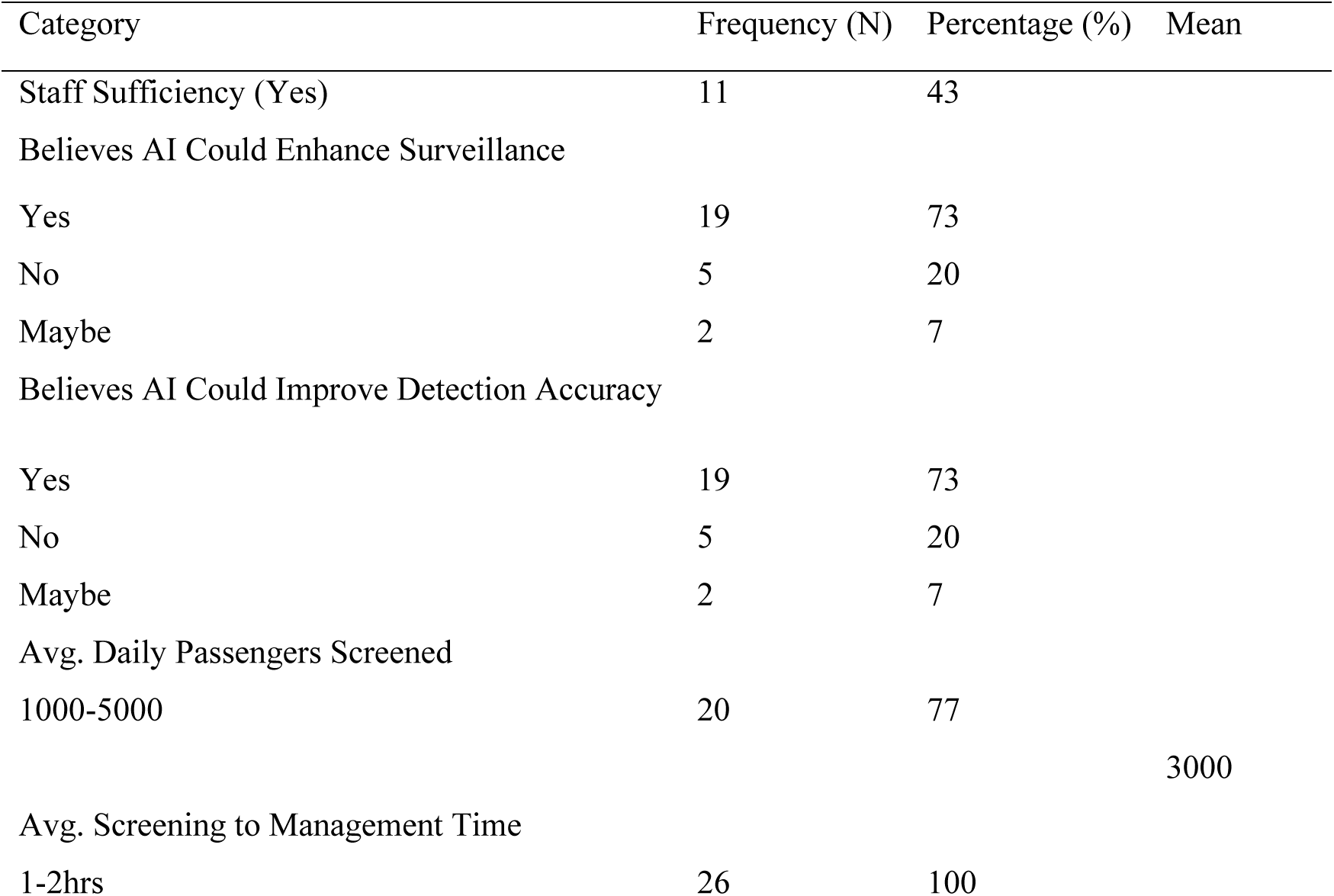
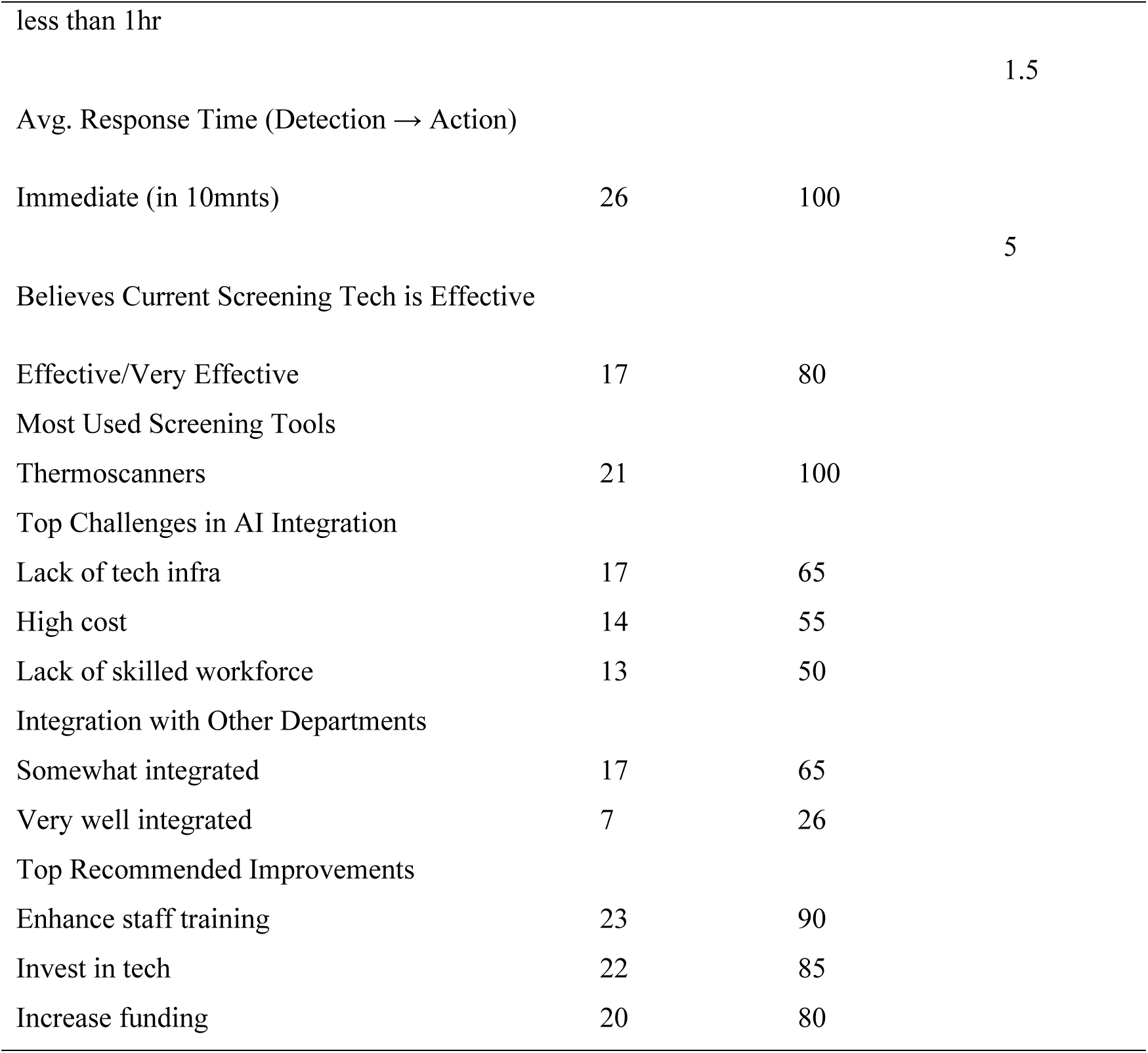
Quantitative analysis of current disease surveillance practices at JNIA and JKIA (n= 25)

### 3.3 Infrastructure Readiness for AI-Driven Disease Surveillance at JNIA

The infrastructure readiness for AI-driven disease surveillance at JNIA was assessed using a standardized checklist of 17 core components, comparing the available quantities to the required quantities for optimal AI operation. Readiness was calculated using the following formula:

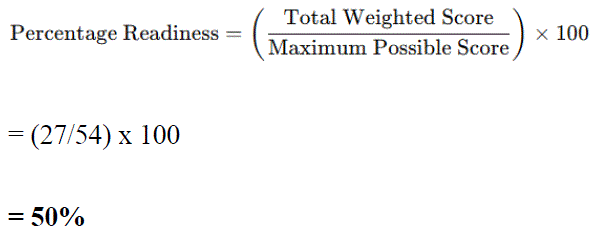

The results showed a total weighted score of 27 out of 54, yielding a 50% readiness score, as shown in Table 3. While the airport has strengths in thermal scanning equipment, AI-based diagnostic systems, and surveillance cameras, critical gaps were identified in AI data processing servers, cloud storage, and automated data transmission systems. Additionally, there was a lack of physical infrastructure for housing AI systems and a shortage of trained AI personnel and health experts to oversee systems.

**Table 3:** Infrastructure readiness checklist at JNIA.

| Sn | Infrastructure | Existing<br>status<br>(1/0) | Available<br>quantity | Needed<br>Quantity |
| --- | --- | --- | --- | --- |
| 1 | AI-based diagnostic systems (such as fever and rash detection modules) | 0 | 0 | 3 |
| 2 | Surveillance cameras with high-resolution image capture | 1 | 6 | 10 |
| 3 | Thermal scanning equipment | 1 | 6 | 5 |
| 4 | High-speed internet connection | 1 | 1 | 1 |
| 5 | Data storage and cloud computing infrastructure | 0 | 0 | 2 |
| 6 | Power backup systems (e.g., generators, UPS) | 1 | 3 | 3 |
| 7 | Artificial Intelligence (AI) data processing servers | 0 | 0 | 4 |
| 8 | Automated data transmission systems (e.g., wireless networks, Bluetooth, or other IoT devices) | 0 | 0 | 2 |
| 9 | Facial recognition or biometric scanning system | 1 | 3 | 1 |
| 10 | Passenger health screening points (e.g., designated areas for testing, monitoring, and diagnosis) | 1 | 3 | 3 |
| 11 | Physical infrastructure for AI system housing (e.g., server rooms, data centers) | 0 | 0 | 2 |
| 12 | Trained AI and data science personnel | 0 | 0 | 5 |
| 13 | Health personnel for system oversight (public health experts) | 0 | 0 | 6 |

| Sn | Infrastructure | Existing status (1/0) | Available quantity | Needed Quantity |
| --- | --- | --- | --- | --- |
| 14 | Stakeholder engagement platforms (e.g., communication systems with public health authorities) | 1 | 2 | 2 |
| 15 | Emergency medical response systems and facilities (e.g., isolation units, medical supplies) | 1 | 2 | 3 |
| 16 | Airport-wide surveillance infrastructure integration (for example, linking AI systems with airport security systems) | 0 | 0 | 1 |
| 17 | Public health reporting tools for data analysis and decision-making | 1 | 1 | 1 |
|  | Total |  | 27 | 54 |

### 3.4. Sociodemographic Characteristics of Qualitative Respondents

The qualitative interviews at JNIA involved 8 respondents, including representatives from the Ministry of Health (MoH), Port Health Officers, IT personnel, and airport management. Table 4 outlines the gender and age distribution, with 62.5% male and 37.5% female respondents. The age distribution included 25% between 25-35 years, 37.5% between 35-45 years, and 37.5% aged 45 and above. The diverse roles of the respondents contributed to a comprehensive understanding of the current disease surveillance operations and the potential challenges for AI adoption at JNIA.

**Table 4:**
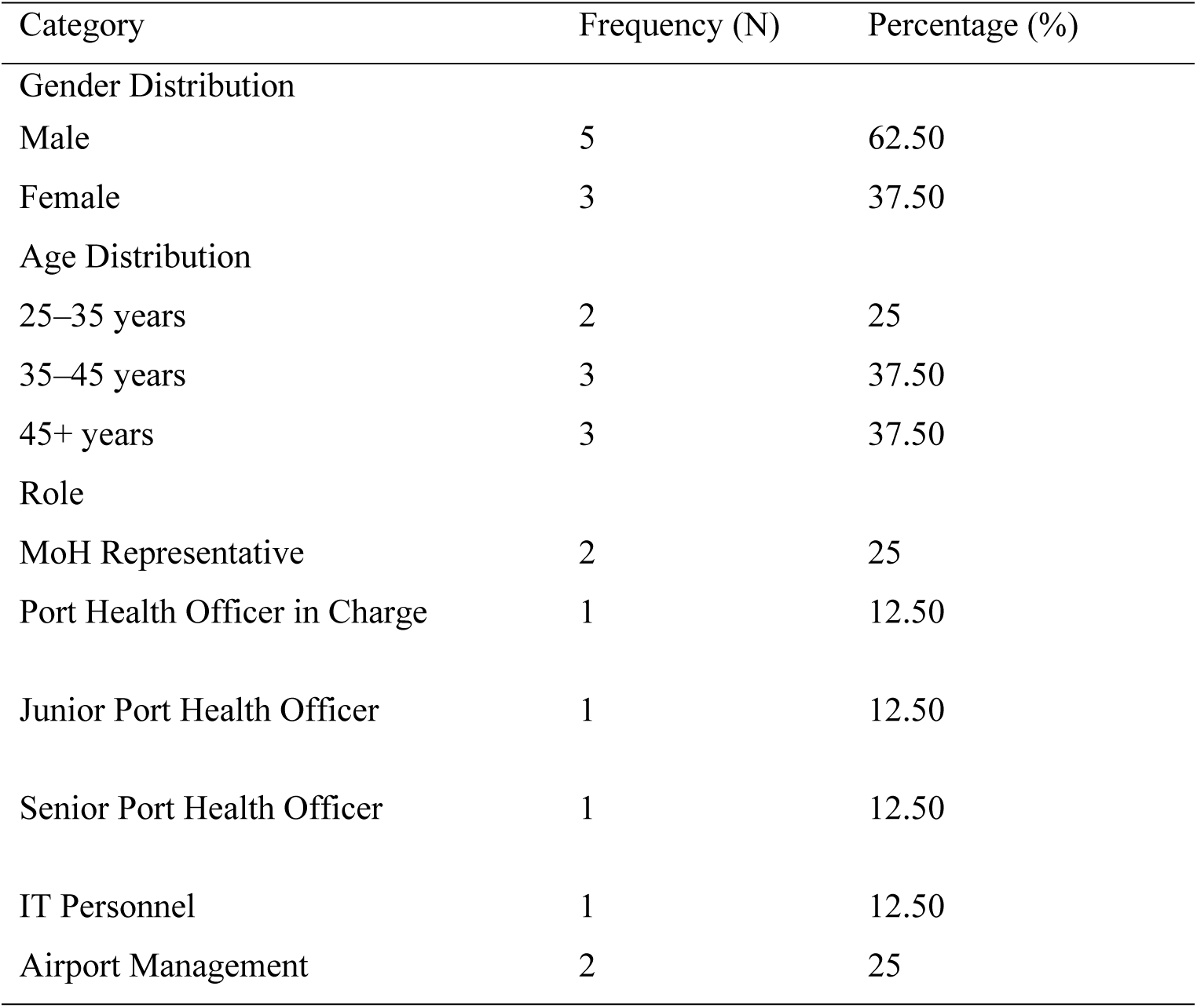
Sociodemographic characteristics of qualitative respondents (n=8)

### 3.5. Thematic Presentation of Qualitative Findings

The hierarchical code tree below structures findings around three parent themes, each with both deductive and inductive emergent sub-themes, providing a comprehensive understanding of the barriers and facilitators to AI-driven disease surveillance at JNIA.

│

├─ THEME 1: Infrastructure & Resource Capacity (Deductive Base)

│ ├─ 1.1 Digital infrastructure gaps
│ ├─ 1.2 Limited technical resource availability
│ ├─ 1.3 Workforce capacity & readiness constraints
│ └─ 1.4 Inductive emergent: Global institutional investment & external resource support

│

├─ THEME 2: Ethical, Privacy & Socio-Cultural Issues (Deductive Base)

│ ├─ 2.1 Privacy and Data Governance
│ ├─ 2.2 Socio-cultural acceptability & operational fit
│ ├─ 2.3 Inductive emergent: Public Awareness Toward AI Surveillance
│ └─ 2.4 Inductive emergent: Cross-border ethical data sharing norms

│

└─ THEME 3: Policy & Regulatory Frameworks & Governance (Deductive base)

├─ 3.1 Inadequate domestic policy & regulatory provisions
├─ 3.2 Weak inter-agency coordination mechanisms
├─ 3.3 Inductive emergent: Government engagement & regional health cooperation
├─ 3.4 Inductive emergent: Public–private sector collaboration governance
├─ 3.5 Inductive emergent: Political will & leadership for AI public health advocacy
└─ 3.6 Inductive emergent: Regional integration & cross-border data regulatory alignment

#### 3.5.1 Theme 1: Infrastructure & Resource Capacity (Deductive Base)

This theme captures the critical challenges related to infrastructure, technical resources, and workforce readiness for AI integration at JNIA, corroborating quantitative findings on infrastructure gaps and staffing constraints. Four sub-themes emerged, including one inductive emergent sub-theme related to external resource support.

##### Digital Infrastructure Gaps

Stakeholders at JNIA consistently highlighted outdated digital infrastructure as a primary barrier to AI integration. Respondents emphasized that legacy systems lack the computing power and connectivity required for real-time AI applications, which depend on robust data handling capabilities. A senior IT expert at JNIA noted: *“Implementing AI will not be easy due to the outdated systems in place.”* A Senior Port Health Officer further elaborated: *“The technology available is just too slow for real-time AI applications. We need faster data processing speeds to make AI work.”* These qualitative insights align with the quantitative infrastructure assessment, which revealed a 50% readiness score and critical gaps in AI data processing servers, cloud storage, and automated data transmission systems, all of which are essential for AI-driven surveillance.

##### Limited Technical Resource Availability

Respondents emphasized that JNIA lacks the foundational technical resources needed to support AI systems at full capacity. A Port Health Officer noted, *“There is a significant gap in the technology we have. Our systems are not ready to support such advanced technology.”* Another respondent highlighted the need for high-speed networks, sufficient bandwidth, and cloud-based storage, components currently absent at JNIA: *“We don’t have the technological foundation needed to support AI systems at full capacity.”* These concerns mirror quantitative findings, where 65% of PHOs identified a lack of technological infrastructure as the top challenge to AI integration, followed by high costs (55%).

##### Workforce Capacity & Readiness Constraints

Workforce readiness emerged as a key barrier, with respondents noting that current JNIA staff lack AI-specific skills and training. An Airport Operations manager stated, *“There may be a reduction in staffing requirements, but the workforce will need reskilling.”* A Ministry of Health (MoH) representative emphasized: *“The workforce isn’t trained to handle such advanced technology, so we need to invest heavily in training.”* Another respondent added that reskilling should include data literacy: *“We need to teach the workforce how to interpret the data AI systems provide and make decisions accordingly.”* Quantitative data support these findings: 50% of PHOs identify a lack of a skilled workforce as a key challenge, and 90% recommend enhanced staff training as a top priority for improvement.

##### Inductive Emergent: Global Institutional Investment & External Resource Support

Stakeholders emphasized that the high costs of AI technology, infrastructure upgrades, and training programs are beyond local capacity. A Senior Port Health Officer noted: *“International organizations such as WHO have a critical role to play in this area. Their support in terms of investment and advocacy can help us develop the necessary infrastructure and regulatory frameworks for the successful deployment of AI systems.”* This aligns with quantitative findings that 80% of PHOs recommended increasing funding as a key improvement, highlighting the need for external support.

#### 3.5.2 Theme 2: Ethical, Privacy & Socio-Cultural Issues (Deductive Base)

This theme focuses on ethical concerns, data privacy, and socio-cultural factors influencing public and stakeholder acceptance of AI-driven surveillance at JNIA. Four sub-themes emerged, including two inductive emergent sub-themes related to public awareness and cross-border data sharing.

##### Privacy and Data Governance

Respondents at JNIA emphasized the sensitive nature of health data and the need for robust data protection protocols to prevent misuse. A senior IT expert noted: *“Passengers may hesitate due to concerns about data privacy and misuse.”* A Senior Port Health Officer stressed the importance of compliance with local and international data protection standards: *“Data Privacy and Protection: Ensure that personal information is not misused by surveillance systems.”* Transparency in data usage was also highlighted as critical for building public trust, with one respondent stating: *“Transparency is key. We need to clearly communicate how data will be used to build trust with the public.”* These concerns reflect the need for clear data governance frameworks, which are currently lacking at JNIA, as noted in the policy and regulatory theme below.

##### Socio-Cultural Acceptability & Operational Fit

Stakeholders emphasized that AI systems must be culturally appropriate and aligned with local privacy expectations to gain public acceptance. A MoH representative stated: *“When we implement AI systems, we must ensure they are culturally appropriate, not just technologically advanced.”* Respondents noted that privacy concerns in East African contexts may differ from those in Western countries, and AI systems must be adapted to these cultural norms to avoid resistance.

##### Inductive Emergent: Public Awareness Toward AI Surveillance

An emergent sub-theme was the critical role of public awareness in ensuring AI adoption. Respondents noted that public understanding of AI’s benefits and data privacy practices is essential to overcoming resistance. Stakeholders recommended targeted public education campaigns to alleviate privacy concerns and promote the value of AI in improving disease surveillance, emphasizing that cultural sensitivity is key to resonating with local communities.

##### Inductive Emergent: Cross-Border Ethical Data Sharing Norms

Respondents discussed the importance of cross-border data sharing for effective AI-driven surveillance, given the regional nature of disease outbreaks. However, they also identified geopolitical challenges, policy inconsistencies, and trust issues between neighboring countries as potential barriers. An Airport Management representative noted: *“Geopolitical issues and differences between neighboring countries hinder the potential for collaboration in disease surveillance. Overcoming these will require regional agreements and trust-building measures.”*

#### 3.5.3 Theme 3: Policy & Regulatory Frameworks & Governance (Deductive Base)

This theme addresses gaps in policy, regulation, and governance that impede AI integration at JNIA. Six sub-themes emerged, including four inductive emergent sub-themes: regional cooperation, public-private partnerships, political leadership, and cross-border regulatory alignment.

##### Inadequate Domestic Policy & Regulatory Provisions

Stakeholders at JNIA consistently highlighted the lack of AI-specific regulatory frameworks in Tanzania. While general data protection laws exist, there are no guidelines tailored to AI implementation in airport port health contexts. A Senior Port Health Officer noted: *“There is a need for clearer regulations that govern AI in disease surveillance.”* Another respondent added: *“Current government policy lacks clear guidelines for implementing AI surveillance.”* Respondents emphasized that existing laws are outdated and do not address the complexities of AI in health surveillance, creating uncertainty for stakeholders and delaying adoption.

##### Weak Inter-Agency Coordination Mechanisms

Respondents noted that effective AI integration requires collaboration across multiple agencies, including health, transport, and IT authorities. However, current governance structures lack provisions for fostering this coordination, leading to fragmented efforts. A Senior MoH representative stated: *“The coordination between different agencies involved in health surveillance is not as strong as it should be. AI adoption needs collaboration across ministries and departments.”* This is consistent with quantitative data showing that only 26% of PHOs reported very good integration between port health and other airport departments.

##### Inductive Emergent: Government Engagement & Regional Health Cooperation

An emergent sub-theme was the need for stronger government engagement in enabling AI adoption at JNIA. Respondents emphasized that the Tanzanian government should play a proactive role in developing regional health policies and forging cross-border partnerships. One Operations department representative noted: *“To successfully implement AI surveillance, the government needs to actively participate in creating regional health policies and forging partnerships with neighboring countries for collaborative data sharing.”*

##### Inductive Emergent: Public-Private Sector Collaboration Governance

Stakeholders highlighted the lack of public-private partnerships (PPPs) as a key policy gap. The absence of collaboration between the public sector and AI technology providers hinders innovation and resource mobilization. A Senior MoH representative stated: *“There is a significant gap between public sector goals and private sector capabilities. A public-private partnership is essential to fast-track AI adoption in disease surveillance.”*

##### Inductive Emergent: Political Will & Leadership for AI Public Health Advocacy

Respondents called for more proactive political leadership in Tanzania to drive AI adoption in public health. They noted that strong political will is needed to secure resources, develop AI-specific policies, and advocate for AI integration in port health. One respondent noted, *“Without political leadership, the potential of AI-driven surveillance remains limited”*.

##### Inductive Emergent: Regional Integration & Cross-Border Data Regulatory Alignment

Respondents emphasized that AI-driven surveillance can only be fully effective with aligned regulatory frameworks across East Africa to facilitate cross-border data sharing. Current policies in Tanzania are inadequate for fostering this alignment, and a unified regional policy governing data-sharing protocols is needed. Stakeholders noted that harmonizing data privacy and security standards across countries would enable seamless AI surveillance and swift response to regional disease outbreaks.

### 3.6. Summary of key Barriers and Facilitators at JNIA

At Julius Nyerere International Airport (JNIA), the integration of AI-driven disease surveillance is influenced by various barriers and facilitators. These are categorized according to three core themes: Infrastructure & Resource Capacity, Ethical, Privacy & Socio-Cultural Issues, and Policy, Regulatory & Governance.

#### 3.6.1. Theme 1: Infrastructure & Resource Capacity

##### Barriers

Staffing Limitations: A significant barrier at JNIA is the shortage of Port Health Officers (PHOs), with only 43% reporting adequate staffing levels. This shortage is especially pronounced in the areas of trained AI/data science personnel and health staff for oversight. The shortage of trained personnel directly affects the efficiency of disease surveillance processes, including long screening-to-management times that average 1.5 hours. This delay undermines the real-time capabilities that AI-driven systems could offer.

Infrastructure Gaps: Critical infrastructure gaps at JNIA limit the deployment of AI technologies in disease surveillance. There is a lack of essential components such as cloud storage, AI data-processing servers, and automated data transmission systems, all of which are necessary for AI systems to process data in real time. The absence of physical infrastructure for housing AI systems, such as server rooms or data centers, also impedes progress. Outdated legacy systems further hinder the integration of AI applications, as they cannot support the data-heavy requirements of AI-driven disease surveillance.

##### Facilitators

Staff Training: To address staffing limitations, it is essential to invest in structured staff training programs focused on AI technologies and data science. Equipping Port Health Officers with the necessary skills to manage and operate AI systems will help alleviate existing challenges and improve operational efficiency. Additionally, enhancing the existing surveillance technology through investments in modern diagnostic tools will expedite screening workflows and reduce pressure on staff.

Infrastructure Upgrades: Upgrading JNIA’s infrastructure is crucial for improving AI readiness. This includes investing in cloud storage solutions, AI data-processing servers, and automated transmission systems. Creating dedicated physical infrastructure, such as server rooms or data centers, will be key to housing AI systems and enabling their successful deployment. Modernizing existing systems to accommodate AI technologies is also essential for optimizing real-time disease surveillance capabilities.

#### 3.6.2. Theme 2: Ethical, Privacy & Socio-Cultural Issues

##### Barriers

Ethical and Privacy Concerns: Ethical concerns, particularly regarding data privacy, pose significant barriers to the adoption of AI-driven disease surveillance at JNIA. There is widespread fear among both airport staff and the public that sensitive health data could be misused. This is compounded by a lack of public awareness of AI surveillance’s benefits, which exacerbates resistance to AI adoption. Without clear data privacy protocols and transparent communication about AI use, these concerns will persist, hindering the acceptance and implementation of AI technologies.

##### Facilitators

Public Awareness Campaigns: Public education campaigns are essential to overcoming privacy concerns and fostering trust in AI surveillance systems. These campaigns should focus on the benefits of AI for enhancing disease detection and public health while addressing privacy and data security concerns. Clear, transparent communication on how health data will be used, safeguarded, and shared will help build public trust and increase acceptance of AI technologies. Additionally, ensuring that the campaigns are culturally sensitive and tailored to the specific concerns of the East African context will improve their effectiveness in gaining public support.

Designing Culturally Appropriate AI Systems: AI systems must be tailored to JNIA’s socio-cultural context. It is essential to ensure that AI surveillance systems align with local privacy expectations and cultural norms. Designing culturally appropriate systems will help reduce resistance and increase the likelihood of public acceptance. Ensuring that AI applications are not perceived as invasive and are instead seen as beneficial to public health is key to mitigating privacy concerns.

#### 3.6.3. Theme 3: Policy, Regulatory & Governance

##### Barriers

Regulatory and Governance Challenges: A lack of AI-specific regulatory policies at JNIA represents a significant barrier to AI adoption. Existing regulations do not address the unique challenges posed by AI technologies in disease surveillance, particularly concerning data governance, system integration, and security protocols. This policy gap creates uncertainty and delays the effective implementation of AI systems. Weak inter-agency coordination and a lack of political will to prioritize AI integration further complicate matters. Moreover, geopolitical barriers related to cross-border data sharing undermine the ability to integrate AI systems effectively, as regional collaboration is essential for comprehensive disease surveillance.

##### Facilitators

Policy Development: To overcome regulatory barriers, JNIA must develop clear, national AI-specific regulatory guidelines for disease surveillance. These guidelines should address critical aspects such as data privacy, ethical considerations, and the integration of AI systems. Strengthening political will and governmental engagement in AI adoption will be essential for advancing these policies. Additionally, fostering regional cooperation on AI policies and developing cross-border data-sharing agreements will enable seamless data exchange and facilitate the integration of AI-driven surveillance systems across East Africa.

Strengthening Inter-Agency Coordination: Effective AI integration at JNIA requires enhanced coordination among relevant government agencies, including health, transport, and IT. Strengthening inter-agency collaboration will ensure that all stakeholders are aligned and working together towards the successful implementation of AI technologies. Establishing clear communication and coordination platforms among these agencies will help streamline AI adoption and address potential conflicts or misunderstandings.

## 4. Discussion

### 4.1 Sociodemographic Characteristics and Implications for AI Adoption

The sociodemographic profile of Port Health Officers (PHOs) at JNIA reveals both opportunities and challenges for AI adoption. With 67% of the workforce aged 25-45 years, the majority of officers fall within a youthful demographic, which is often associated with higher digital literacy and greater adaptability to new technologies. This is consistent with findings from other African studies, which suggest that younger workforces tend to embrace digital health solutions more readily (9). At JNIA, the average age of 40 years indicates a workforce that can be trained to acquire new technical competencies, particularly those related to AI integration. However, the gender disparity at JNIA, where 68% of the workforce is male, suggests potential challenges in achieving gender-inclusive AI adoption. Gender-balanced teams in healthcare settings have been shown to be more receptive to collaborative digital solutions(10). The higher proportion of junior officers at JNIA (53.85%) and a relatively longer average employment duration (5 years) suggest that JNIA may benefit from embedding AI competencies early in new staff’s career trajectories. The finding that 15-20% of officers have less than 1 year of experience presents an opportunity to integrate AI training into their foundational skills development, aligning with the WHO Global Strategy on Digital Health 2020-2025, which emphasizes incorporating digital health competencies into pre-service training for health workers in LMICs(11).

### 4.2 Current Disease Surveillance Operations

A significant barrier to integrating AI into disease surveillance at JNIA is insufficient staffing. Only 43% of Port Health Officers (PHOs) report adequate staffing levels, which is problematic considering the demands of high-traffic airports. Effective AI implementation requires not only more personnel but also a trained workforce to manage and operate these systems. In LMICs, workforce capacity has been identified as a limiting factor in the adoption of advanced public health technologies(12). Therefore, targeted training programs that focus on AI, data management, and system oversight are crucial for overcoming this gap. Despite staffing challenges, 73% of officers at JNIA express optimism about AI’s potential to enhance disease surveillance and detection accuracy. This aligns with global findings that AI has proven effective in outbreak prediction, diagnostic accuracy, and real-time disease monitoring (13). The ability of AI to process large datasets and identify patterns rapidly is critical in an airport setting like JNIA, where quick disease detection is essential. However, AI adoption is hindered by insufficient technological infrastructure and high costs. JNIA screens between 1,000 and 5,000 passengers daily, a volume that requires efficient and rapid disease-detection methods. Currently, the average screening-to-management time is 1.5 hours, which is notably slower than best practice benchmarks. AI integration could streamline this process by automating data collection and analysis, ultimately reducing response times. A lack of strong inter-departmental integration, with only 26% of officers reporting well-integrated systems, further compounds the efficiency issue. Effective disease surveillance requires seamless collaboration among health, security, and immigration departments (14). Current screening technologies, including thermoscanners, are rated as effective by 80% of PHOs. However, these tools are limited in detecting asymptomatic or mildly symptomatic passengers. AI-enhanced systems that integrate thermal data with additional health information, such as travel history and health declarations, could improve detection specificity and sensitivity, thereby enhancing the comprehensiveness of disease surveillance(15). AI integration faces several key barriers, including outdated technological infrastructure (65%), high costs (55%), and a lack of skilled personnel (50%). These challenges are common across LMICs, where initial investments in AI systems are often prohibitive(16). Additionally, AI requires specialized skills for system operation and data interpretation, a gap that can only be addressed through targeted training programs for existing personnel. Inter-departmental collaboration remains a significant challenge. Successful AI-driven systems rely on data sharing across various airport departments. Without this integration, AI tools cannot access the comprehensive datasets necessary for accurate predictions and timely alerts. Research underscores the importance of cross-agency collaboration for the successful integration of AI in public health surveillance(14). JNIA’s PHOs have identified key priorities to improve disease surveillance, including enhanced staff training (90%), investment in technology (85%), and increased funding (80%). These recommendations align with the WHO Global Strategy on Digital Health, which emphasizes workforce capacity-building, infrastructure investment, and sustainable financing as foundational pillars for digital health transformation(11). Prioritizing training and infrastructure development will be crucial to enabling JNIA to successfully leverage AI for improved disease surveillance.

### 4.3 Infrastructure Readiness: Partial Strengths and Critical Gaps for AI Integration

The integration of AI into disease surveillance at JNIA is significantly hindered by infrastructure gaps. With a 50% readiness score, JNIA has basic surveillance capabilities, such as thermal scanners and surveillance cameras, but lacks critical AI-specific infrastructure. These include essential components such as AI data-processing servers, cloud storage, and automated data transmission systems, all needed to process and analyze large datasets in real time. JNIA’s existing infrastructure offers opportunities for incremental upgrades, particularly in enhancing surveillance cameras with AI-powered video analytics to detect additional symptoms, such as coughs or rashes, and integrating AI algorithms into thermal scanners to improve detection accuracy. Power backup systems, which meet the required standards, are vital to ensure AI systems can function reliably, even during power interruptions, a crucial consideration in LMICs. However, significant infrastructure gaps remain. JNIA does not have cloud-based storage or AI data-processing servers, limiting its ability to store and process large volumes of health data. Without cloud storage, the AI systems would be confined to on-premises solutions that are less efficient and more costly. Additionally, there is no physical infrastructure, such as dedicated server rooms or data centers, to house AI systems securely, which is essential for maintaining system integrity and preventing overheating or data breaches. Workforce limitations further compound these challenges, as JNIA lacks trained AI and data science personnel. Effective AI integration requires specialized skills for system monitoring and data interpretation, which are currently lacking. To bridge this gap, targeted training programs should focus on building data literacy and AI system operations, particularly for PHOs. Another critical barrier is the lack of integration between disease surveillance systems and other airport-wide infrastructure. For AI systems to be effective, seamless data sharing across airport departments, such as immigration and security, is essential. JNIA must prioritize establishing clear data-sharing protocols to improve the feasibility of AI systems and overall surveillance efficiency.

### 4.4 Thematic Domains

The study’s three core thematic domains, Infrastructure & Resource Capacity, Ethical/Privacy/Socio-Cultural Issues, and Policy & Regulatory Frameworks, provide a comprehensive framework for assessing AI feasibility at JNIA. Below is a complete analysis of each theme.

#### 5.4.1 Theme 1: Infrastructure & Resource Capacity

The infrastructure and resource capacity at JNIA reveal significant challenges to implementing AI-driven disease surveillance. According to a senior IT expert at JNIA, outdated systems, referred to as “technological lock-in”, hinder AI adoption, as manual processes and legacy hardware cannot support the real-time analytics required by AI. These deficiencies are consistent with challenges identified globally in LMICs, where AI adoption is often impeded by similar infrastructure gaps(17).

The absence of cloud infrastructure is particularly significant, as cloud-based systems enable scalable data storage and efficient processing, which are essential for AI to handle large datasets. Without such infrastructure, JNIA’s AI applications would be limited to on-premises solutions that are less efficient, more costly, and unable to integrate data across different surveillance systems. This gap, compounded by slow processing speeds, further restricts AI’s potential. As one Port Health Officer (PHO) noted, *“the existing technology is too slow for real-time AI applications, limiting AI systems’ ability to process data at the required speed to maintain passenger flow and detection accuracy”.* The challenges go beyond just hardware. JNIA’s workforce faces significant limitations, as no AI-specific training has been provided, and staff lack the specialized skills required to operate and interpret AI systems. This reflects a broader trend in LMICs, where healthcare workers are typically trained in clinical practice rather than in digital technologies (18). As a result, there is a pressing need for workforce reskilling to ensure that Port Health Officers can transition from manual screening processes to oversight of AI systems. This shift involves training in data literacy, system monitoring, and the interpretation of algorithmic outputs, which are essential for making informed decisions based on AI-generated insights. Furthermore, there is a notable gap in technical support and maintenance. As one PHO observed, *“We don’t have the technological foundation needed to support AI systems at full capacity.”* This lack of in-house expertise in AI maintenance is a common issue in LMICs, where the cost and delayed response of external technical support pose a risk to the long-term sustainability of AI systems(19). For JNIA, this means that any AI system deployed would be vulnerable to operational failures unless paired with robust, locally available technical training and support. An emerging theme identified in the qualitative interviews is the critical role of global institutional investment. JNIA stakeholders repeatedly emphasized that local resource constraints make the integration of AI unfeasible without external funding and technical support. This aligns with broader literature findings that AI adoption in public health settings, particularly in LMICs, depends heavily on international partnerships, such as collaborations with global health organizations and technology firms(20). Without these external resources, JNIA’s current infrastructure and workforce limitations will continue to hinder AI integration. A systemic approach involving infrastructure upgrades, targeted workforce reskilling, and external funding will be necessary to overcome these barriers and enable the effective deployment of AI-driven disease surveillance at JNIA.

#### 5.4.2 Theme 2: Ethical, Privacy, and Socio-Cultural Issues

Ethical concerns, especially regarding privacy and data security, are major obstacles to AI adoption at Julius Nyerere International Airport (JNIA). The collection and processing of sensitive passenger health data by AI systems pose significant privacy risks, including unauthorized access, data breaches, and potential misuse. One PHO expressed concerns about the increased volume of data AI would collect, emphasizing the need for robust data protection measures. This aligns with global standards that emphasize data privacy as essential to maintaining public trust in digital health systems (21). In LMICs, this challenge is intensified by weak data protection frameworks, underscoring the need for JNIA to incorporate privacy-by-design principles into AI systems. These should include measures such as data anonymization, limited access controls, and encryption of sensitive data. Algorithmic bias and fairness also emerged as key ethical issues. AI systems, often trained on historical data, may perpetuate biases that affect certain demographic groups, leading to inaccurate disease detection or unfair outcomes. For instance, if training data underrepresents specific groups, AI systems might perform poorly in detecting diseases in these populations, exacerbating health inequities. An IT expert emphasized the need for bias mitigation strategies, including diverse training datasets, regular algorithmic audits, and a human-in-the-loop approach in which PHOs review AI outputs to ensure fairness. Addressing these biases is essential to ensure equitable and accurate disease surveillance. Socio-cultural factors play a significant role in AI adoption at JNIA. Public trust in digital health systems is shaped by local cultural norms around privacy and interpersonal interactions. As noted by a JNIA stakeholder, *“passengers may prefer interacting with human officers rather than AI systems due to concerns about data misuse.”* This preference is common in East Africa, where face-to-face consultations are valued in healthcare(22). Additionally, low digital literacy among passengers may hinder acceptance of AI, as some may fear that it could misinterpret their health data. Public awareness campaigns that explain AI’s role in disease surveillance, how data is protected, and its benefits are essential to building trust and acceptance. Concerns about workforce autonomy and job security were also raised, with some PHOs fearing that AI might replace their roles. However, AI is more likely to augment human expertise, automating routine tasks and freeing up staff for more complex decision-making. To alleviate these concerns, JNIA must ensure transparent communication, guarantee job security, and offer training programs that position AI as a tool for enhancing, not replacing, human capacity. Addressing the ethical, privacy, and socio-cultural concerns surrounding AI integration at JNIA requires a comprehensive approach. This should integrate privacy safeguards, bias mitigation strategies, public trust-building, and workforce engagement to ensure that AI systems are not only technically viable but also ethically sound and socially acceptable.

#### 5.4.3 Theme 3: Policy & Regulatory Frameworks

The integration of AI at JNIA is significantly hindered by fragmented policy and regulatory frameworks. Tanzania’s digital health policies, such as the National Digital Health Strategy (2021-2025), focus on general health transformations but lack specific provisions for AI in public health or airport surveillance(23). This absence of AI-specific regulations creates uncertainty about key issues such as data governance, algorithmic accountability, and compliance with international health standards like the WHO’s International Health Regulations (IHR 2005), which require robust disease surveillance and data sharing. The lack of clear guidelines on AI system design, data collection, and algorithm transparency poses risks to JNIA’s management of passenger health data. Without explicit regulations governing how AI should collect, store, and share this data, particularly across borders, JNIA’s ability to meet both national and international data protection standards is compromised. This gap is particularly critical for international airports, where cross-border data sharing is essential for detecting global health threats. Tanzania’s existing data protection laws provide a basic framework for privacy, but do not address the complexities of AI-driven data processing. For instance, the Act does not cover issues such as automated decision-making or the sharing of health data across countries, which are necessary for AI systems at international airports. Further complicating AI adoption at JNIA is the lack of alignment between Tanzania’s national health regulations and AI’s potential to enhance disease surveillance. While AI could support Tanzania’s compliance with the IHR by improving the detection and reporting of health threats, there are no clear guidelines connecting AI adoption to IHR compliance. This regulatory gap represents a missed opportunity to harness AI’s capabilities in global health surveillance. Additionally, institutional coordination between JNIA and the Tanzanian Ministry of Health is lacking, with no formal mechanism for aligning AI initiatives with national health priorities. This lack of coordination prevents JNIA from accessing national resources for AI training and infrastructure, and hinders the integration of AI into broader national health efforts. Finally, the lack of clear national funding mechanisms for AI in public health means JNIA must rely on external donors or limited internal budgets, which are often inconsistent and short-term. This funding uncertainty threatens the sustainability of AI initiatives at JNIA and reflects broader challenges LMICs face in securing long-term financing for digital health systems. To address these policy and regulatory gaps, it is crucial for Tanzania to update its digital health policies to include AI-specific provisions, aligning with international guidelines such as the WHO’s AI for Health Guidelines. JNIA should also develop internal protocols for AI implementation, including data governance and algorithmic accountability, and establish a coordination mechanism with the Ministry of Health to ensure alignment with national health priorities. The fragmented policy and regulatory landscape at JNIA presents a significant barrier to the effective integration of AI in disease surveillance. Addressing these gaps is essential to ensure AI adoption is legally compliant, sustainable, and aligned with global health regulations.

### 5.7 Site-Specific Barriers and Facilitators at JNIA

The study emphasizes key barriers to AI adoption at JNIA, including staffing shortages, outdated infrastructure, a lack of AI-specific policies, and weak coordination between agencies. However, important enablers like a young workforce, staff optimism about AI, and existing surveillance technologies offer a foundation for future AI integration. Overcoming these challenges through focused efforts in workforce training, infrastructure upgrades, and policy development is crucial for advancing AI adoption.

### 5.8 Study Contributions

The study offers critical insights into the feasibility of AI-driven disease surveillance at JNIA. It contributes to the global literature by contextualizing the challenges and opportunities of AI in LMICs, particularly in East African airports, and by highlighting the need for region-specific solutions to address infrastructure gaps, staffing limitations, and regulatory challenges.

#### Study Limitations and Future research

This study, while providing valuable insights into the feasibility of AI at JNIA, has several limitations. The relatively small sample size, comprising 26 PHOs and 8 key informants, limits the diversity of perspectives, particularly for qualitative data, and introduces potential bias due to self-reported data. Additionally, the study’s cross-sectional design captures only a snapshot of the current state, without considering longitudinal changes in infrastructure, policy, or workforce capacity. The absence of a formal cost-effectiveness analysis further restricts the ability to assess the financial feasibility and long-term economic benefits of AI-driven disease surveillance. Furthermore, the study focused primarily on health system stakeholders, excluding passenger and public perspectives. This gap limits our understanding of public attitudes toward AI surveillance and privacy concerns, both of which are crucial to the successful implementation of these systems.

Future studies should address these limitations by expanding the sample size to include a broader range of stakeholders, such as passengers and policymakers, and conducting longitudinal research to track the impact of AI integration over time. Additionally, a formal cost-effectiveness analysis would provide policymakers with critical insights into the financial viability of AI adoption. Future research could also incorporate public opinion through surveys or focus groups, which would be essential to comprehensively understand the challenges and opportunities of AI-driven disease surveillance in the region.

### 5.1 Conclusion

This study demonstrates that implementing AI-driven systems for early detection of infectious diseases at JNIA is currently not feasible. This infeasibility arises from several interconnected, systemic gaps that must be addressed before AI integration can be considered viable. These challenges include outdated and fragmented technological infrastructure that currently lacks critical components, such as AI data-processing servers and cloud storage. Additionally, the workforce at JNIA lacks the specialized AI and data science skills necessary to effectively operate and oversee such advanced systems. There is also a lack of tailored regulatory frameworks that address the unique challenges posed by AI technologies, particularly in data governance and ethical deployment. Finally, concerns around data privacy, cultural sensitivity, and public trust in AI surveillance systems remain unresolved, further hindering the adoption of AI technologies.

Although the current conditions make AI adoption unfeasible, the study suggests that with strategic investments in infrastructure, comprehensive workforce training, and the development of clear, AI-specific regulatory frameworks, the integration of AI systems could become feasible in the future. Additionally, regional cooperation and cross-border data-sharing agreements will be vital to overcoming current limitations, enabling AI-driven disease surveillance and enhancing health security. Improving East Africa’s health security is essential not only for the region but also for the world, as it directly supports the global health security agenda. The recommended implementation framework for the future is shown below.

**Figure 2:**
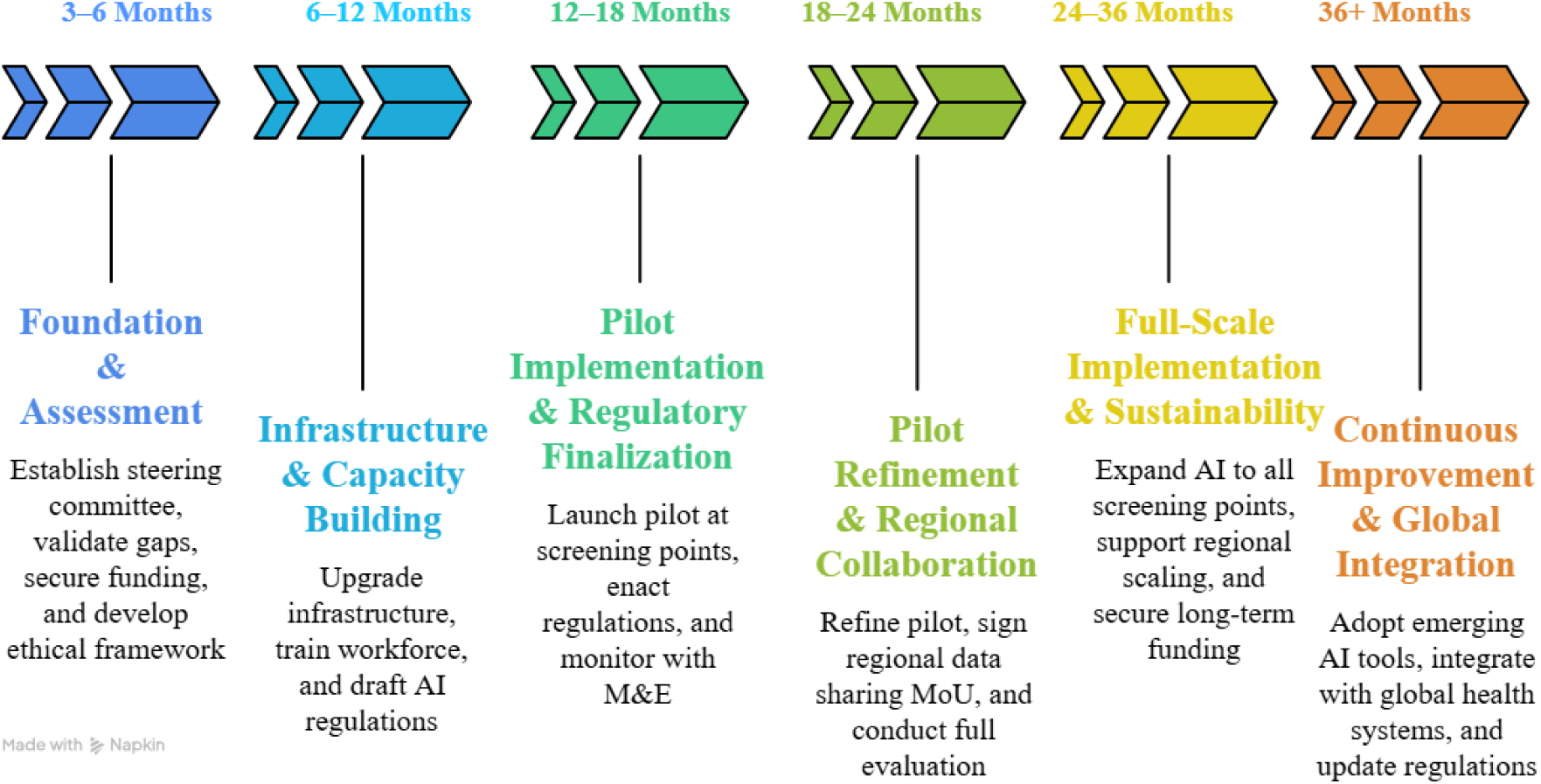
Recommended Implementation Framework for an AI-Driven system for early detection of diseases at International Airports in East Africa

## Acknowledgments

First and foremost, gratitude is extended to God for His guidance and strength throughout this journey. Appreciation is also given to the Ministry of Commerce of the People’s Republic of China (MOFCOM) for their generous scholarship, which made this research possible. Thanks are due to Southern Medical University for providing an inspiring academic environment. Special thanks go to my supervisor, Qun HE, for his invaluable guidance and support throughout this study. Heartfelt thanks are offered to friends and family for their unwavering encouragement and support. Special thanks to my beloved son, Ezekiel, and my lovely mom, Bernadetha Malingumu, for their constant encouragement and support.

## Author Contributions

**Elizabeth Malingumu:** Data collection coordination, Conceptualization, and writing original draft

**Isimbi Badaga:** Validation and Data curation

**Dotto Kisendi:** Methodology, Formal Data analysis and Visualization

**Rasabs Wende Pierre Kabore:** Result arrangement and discussion

**Onacis Guerde Yeremon:** Qualitative data analysis and discussion

**Mohamed Mohamed:** Review, editing, and administration

**Qun HE:** Supervision and Resources

## Data Availability Statement

The data is available from the corresponding author upon reasonable request.

